# Association of Intravenous Bamlanivimab Use with Reduced Hospitalization, Intensive Care Unit Admission, and Mortality in Patients with Mild to Moderate COVID-19

**DOI:** 10.1101/2021.05.23.21257670

**Authors:** Ravindra Ganesh, Colin Pawlowski, John C. O’Horo, Lori L. Arndt, Richard Arndt, Sarah J. Bell, Dennis M. Bierle, Molly Destro Borgen, Sara N. Hanson, Alexander Heyliger, Jennifer J. Larsen, Patrick Lenehan, Robert Orenstein, Arjun Puranik, Leigh L. Speicher, Sidna M. Tulledge-Scheitel, AJ Venkatakrishnan, Caroline G. Wilker, Andrew D. Badley, Raymund R. Razonable

**Affiliations:** Mayo Clinic, Rochester, MN; nFerence, Cambridge, MA; Mayo Clinic Health System, Eau Claire, WI; Mayo Clinic Health System, Mankato, MN; Mayo Clinic in Arizona, Phoenix; Mayo Clinic in Florida, Jacksonville; Mayo Clinic Health System – Franciscan Healthcare, La Crosse, WI

## Abstract

**Background:** Clinical data to support the use of bamlanivimab for the treatment of outpatients with mild to moderate coronavirus disease-19 (COVID-19) is needed.

**Methods:** 2,335 patients who received single-dose bamlanivimab infusion between November 12, 2020 to February 17, 2021 were compared with a propensity-matched control of 2,335 untreated patients with mild to moderate COVID-19 at Mayo Clinic facilities across 4 states. The primary outcome was the rate of hospitalization at days 14, 21 and 28.

**Results:** The median age of the population was 63; 47.3% of the bamlanivimab-treated cohort were ≥65 years; 49.3% were female. High-risk characteristics included hypertension (54.2%), body mass index ≥35 (32.4%), diabetes mellitus (26.5%), chronic lung disease (25.1%), malignancy (16.6%), and renal disease (14.5%). Patients who received bamlanivimab had lower all-cause hospitalization rates at days 14 (1.5% vs 3.5%; Odds Ratio [OR], 0.38), 21 (1.9% vs 3.9%; OR, 0.46), and 28 (2.5% vs 3.9%; OR, 0.61). Secondary exploratory outcomes included lower intensive care unit admission rates at days 14 (0.14% vs 1%; OR, 0.12), 21 (0.25% vs 1%; OR: 0.24) and 28 (0.56% vs 1.1%; OR: 0.52), and lower all-cause mortality at days 14 (0% vs 0.33%), 21 (0.05% vs 0.4%; OR,0.08) and 28 (0.11% vs 0.44%; OR, 0.01). Adverse events were uncommon with bamlanivimab, occurring in 19/2355, most commonly fever (n=6), nausea (n=5), and lightheadedness (n=3).

**Conclusions:** Among high-risk patients with mild to moderate COVID-19, treatment with bamlanivimab was associated with a statistically significant lower rate of hospitalization compared with usual care.

**Funding:** Mayo Clinic.

## Introduction

Control of the Severe Acute Respiratory Syndrome-Coronavirus 2 (SARS-CoV-2) pandemic requires prevention, early diagnosis, and effective treatment. Enhanced testing has allowed for early identification of infected individuals who do not yet require hospitalization but are at high risk for complications. Most treatments authorized or approved in 2020 were targeted at late stage disease and critical illness, but new treatments are emerging to intervene earlier in the course of illness for high-risk individuals. One treatment option for these individuals is anti-spike neutralizing monoclonal antibody, which has been authorized for emergency use by the US Food and Drug Administration (FDA). Bamlanivimab was authorized on November 9, 2020, the combination of casirivimab-imdevimab on November 21, 2020, and the combination of bamlanivimab-etesevimab on February 9, 2021.^1-3^ These monoclonal antibodies inhibit the interaction of SARS-CoV-2 spike protein with ACE-2 receptors, thereby preventing viral attachment and infectivity.^4^ On April 16, 2021, the FDA had revoked the EUA for bamlanivimab monotherapy due to concerns over emerging resistance patterns in SARS-CoV-2 variants.^5^

A phase 2 placebo-controlled trial showed decreased Emergency Department visits and hospitalizations among patients administered bamlanivimab,^6^ leading to it being granted an Emergency Use Authorization (EUA) by the FDA.^1,7^ However, the utilization of bamlanivimab has been slow, owing partly to the complexities of implementing outpatient infusion centers for infectious patients.^8^ Further, some patients were wary of investigational treatment and clinicians were skeptical in recommending these therapies due to limited clinical evidence and initial lack of endorsement by national societies.^9-11^

Mayo Clinic developed dedicated infusion facilities and assembled multidisciplinary teams to coordinate monoclonal antibody infusions to patients with coronavirus disease-19 (COVID-19) who are eligible under the EUA.^12,13^ This study was conducted to analyze the association of bamlanivimab monotherapy with clinical outcomes in high-risk patients with mild to moderate COVID-19.

## Methods

### Monoclonal Antibody Treatment Program

The Mayo Clinic Monoclonal Antibody Treatment (MATRx) program was established on November 7, 2020, and the first patients were infused with bamlanivimab on November 19, 2020. The details of this program have been reported.^12^ A multidisciplinary team reviewed all patients identified from an electronic registry of positive SARS-CoV-2 PCR tests and self- and clinician-referred patients. Under the EUA, patients were eligible for bamlanivimab if they had mild to moderate COVID-19, were within 10 days of symptom onset, and had at least one of the following criteria: age ≥65 years, body mass index (BMI) ≥35, diabetes, chronic kidney disease, immunosuppressive medication use, or an immunocompromising condition. Patients 55 years and older qualified if they had hypertension, cardiovascular disease, or chronic lung disease. All eligible patients were approached by MATRx team members for education and consenting.

### Study Design and Participants

This retrospective study was approved by the Mayo Clinic Institutional Review Board. Informed consent was waived and patients without research authorization were excluded. The study enrolled adult (≥ 18 years old) patients identified from the Mayo Clinic electronic health record (EHR) database with positive SARS-CoV-2 PCR tests between November 12, 2020 and February 17, 2021. The start date November 12, 2020 was selected as it was the earliest test date for a patient who was infused with bamlanivimab monotherapy. The study end date was selected as the most recent date with data available. The participant selection algorithm (**Figure 1**) resulted into two cohorts balanced for relevant demographic and clinical covariates: (1) treated patients who received bamlanivimab infusion, and (2) control patients who did not receive bamlanivimab after COVID-19 diagnosis.

**Figure 1:**
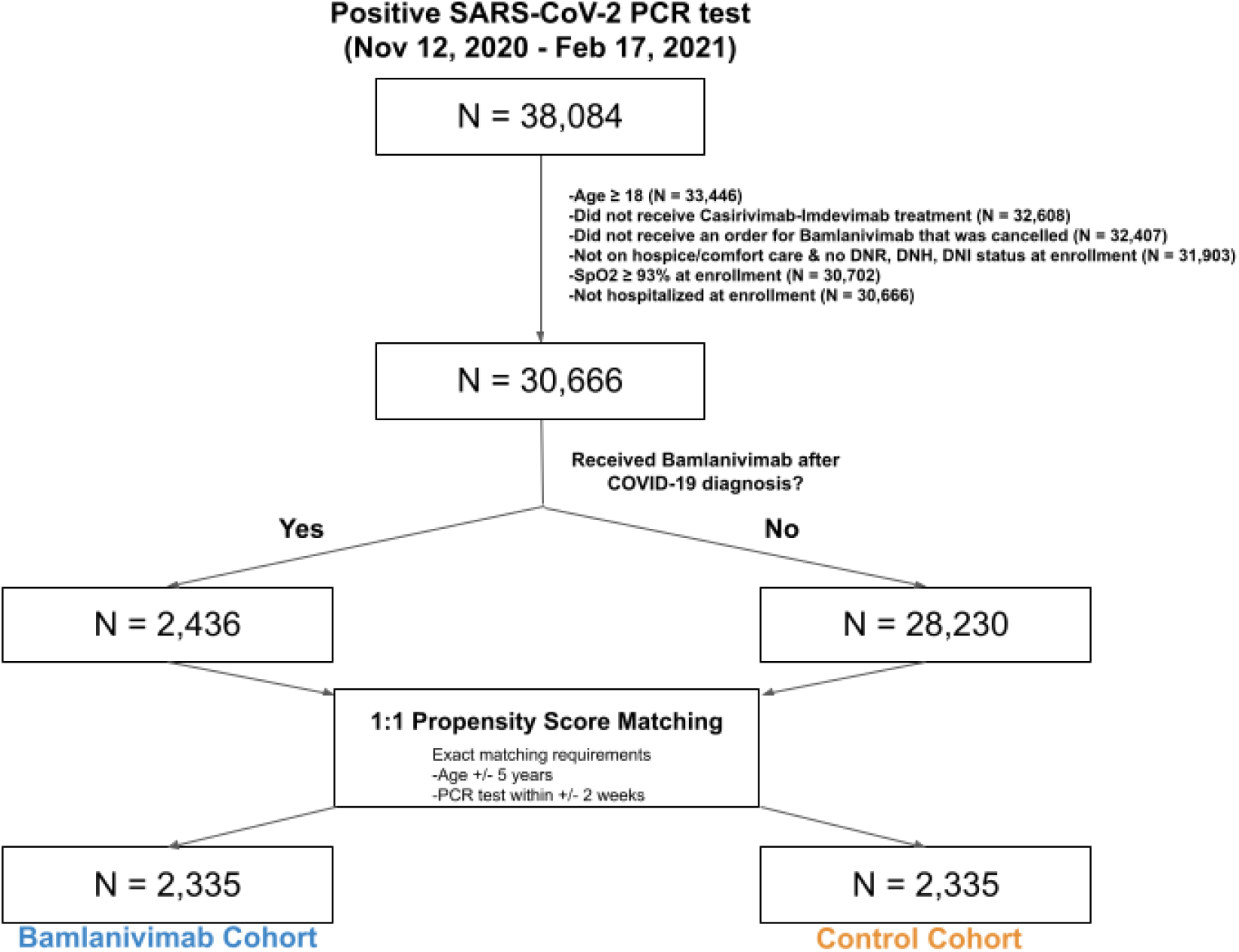
Study Population, Participant Selection and Propensity Matching.

### Participant Selection and Propensity Score Matching

The study population was selected from the pool of adult patients with COVID-19 who met the following criteria: (1) had not received casirivimab and imdevimab at any time during the study period, (2) did not have a cancelled bamlanivimab order, (3) were not on hospice or comfort care, (4) did not have a do not intubate (DNI), do not resuscitate (DNR), or do not hospitalize (DNH) status, (5) had minimum SpO2 of ≥93%, and (6) not currently hospitalized at the time of positive PCR test or bamlanivimab infusion. For each patient in the treated cohort, the enrollment date was defined as the date of bamlanivimab infusion. A histogram of infusion dates relative to PCR diagnosis dates is provided in **eFigure 1**.

Propensity score matching was performed to select matched controls balanced on covariates that may influence bamlanivimab administration (**Table 1**).^15^ Propensity scores were computed for each patient by fitting an L1-regularized logistic regression model to predict which of the two cohorts the patient was in, as a function of the covariates detailed in the next section.^16^ To identify a matched control for each treated patient, a set of control patients with the same age (+/-5 years) and PCR diagnosis date (+/-7 days) was considered, and the patient with the closest propensity score was selected, if the propensity score difference was less than the selected threshold. If the control patient (1) had a minimum SpO2 < 93%, (2) was hospitalized, (3) had an active DNR, DNI or DNH status, (4) was receiving only palliative or comfort care, or (5) was deceased on or before the date of study enrollment, then a new control patient (the next nearest neighbor by propensity score) was selected. Controls could not have received bamlanivimab due to the time limits of the EUA. This process was repeated until an eligible match was found. If an eligible match was not found, the search was expanded to the set of control patients with age +/-5 years and PCR diagnosis dates +/-14 days relative to the bamlanivimab patient. If the expanded search did not find any control patients, the bamlanivimab-treated patient was dropped from the analysis. The caliper threshold was set to 0.1 * pooled standard deviation of the propensity scores in the logit space. For each control patient, the study enrollment date was defined based on the number of days between the positive PCR test and bamlanivimab infusion for the matched treated patient **(eFigure 2**). ^17,18^

**Table 1:** Clinical Characteristics of Bamlanivimab and Control Cohorts.

| Clinical covariate | Before Matching |  |  | After Matching |  |  |
| --- | --- | --- | --- | --- | --- | --- |
|  | Bamlanivimab cohort<br>(n = 2,436) | Control cohort<br>(n = 28,230) | Standardized Difference | Bamlanivimab cohort<br>(n = 2,335) | Control cohort<br>(n = 2,335) | Standardized Difference |
| Age (years) |  |  |  |  |  |  |
| Median | 64 | 44 |  | 63 | 63 |  |
| IQR | (52, 72) | (30, 57) |  | (52, 72) | (52, 71) |  |
| <65 years old | 1,274<br>(52.3%) | 24,566<br>(87.0%) | 0.99 | 1,230<br>(52.7%) | 1,274<br>(54.6%) | 0.04 |
| 65-75 years old | 734 (30.1%) | 2,347<br>(8.3%) | 0.74 | 716 (30.7%) | 662<br>(28.4%) | 0.05 |
| >75 years old | 428 (17.6%) | 1,317<br>(4.7%) | 0.56 | 389 (16.7%) | 399<br>(17.1%) | 0.01 |
| Sex |  |  |  |  |  |  |
| Female | 1201<br>(49.3%) | 14,476<br>(51.3%) | 0.04 | 1,152<br>(49.3%) | 1,154<br>(49.4%) | 0.00 |
| Male | 1235<br>(50.7%) | 13,746<br>(48.7%) | 0.04 | 1,183<br>(50.7%) | 1,181<br>(50.6%) | 0.00 |
| Race |  |  |  |  |  |  |
| American Indian | 4 (0.2%) | 115<br>(0.4%) | 0.04 | 4 (0.2%) | 5 (0.2%) | 0.01 |
| Asian | 37 (1.5%) | 633<br>(2.2%) | 0.05 | 36 (1.5%) | 37 (1.6%) | 0 |
| Black / African American | 49 (2.0%) | 761<br>(2.7%) | 0.04 | 48 (2.1%) | 55 (2.4%) | 0.02 |
| White / Caucasian | 2,270<br>(93.2%) | 23,302<br>(82.5%) | 0.29 | 2,174<br>(93.1%) | 2,164<br>(92.7%) | 0.02 |
| Other<sup>1</sup> | 56 (2.3%) | 1,091<br>(3.9%) | 0.08 | 53 (2.3%) | 64 (2.7%) | 0.03 |
| Unknown | 20 (0.8%) | 2,328<br>(8.2%) | 0.28 | 20 (0.9%) | 10 (0.4%) | 0.05 |
| Ethnicity |  |  |  |  |  |  |
| Hispanic | 121 (5.0%) | 1,969<br>(7.0%) | 0.08 | 114 (4.9%) | 118<br>(5.1%) | 0.01 |
| Non-Hispanic | 2,270<br>(93.2%) | 23,359<br>(82.7%) | 0.28 | 2,176<br>(93.2%) | 2,192<br>(93.9%) | 0.03 |
| Unknown | 45 (1.8%) | 2,902<br>(10.3%) | 0.29 | 45.0 (1.9%) | 25.0<br>(1.1%) | 0.07 |
| Site |  |  |  |  |  |  |
| Scottsdale, Arizona | 353 (14.5%) | 3,790<br>(13.4%) | 0.03 | 342 (14.6%) | 403<br>(17.3%) | 0.07 |
| Jacksonville, Florida | 367 (15.1%) | 2,701<br>(9.6%) | 0.18 | 338 (14.5%) | 401<br>(17.2%) | 0.07 |
| Mayo Clinic Health Systems | 1,217<br>(50.0%) | 17,531<br>(62.1%) | 0.25 | 1,191<br>(51.0%) | 1,115<br>(47.8%) | 0.07 |
| Rochester, Minnesota | 499 (20.5%) | 4,151<br>(14.7%) | 0.16 | 464 (19.9%) | 416<br>(17.8%) | 0.05 |
| BMI (kg/m<sup>2</sup>) |  |  |  |  |  |  |
| Underweight (< 18.5) | 7 (0.3%) | 116<br>(0.4%) | 0.02 | 6 (0.3%) | 10 (0.4%) | 0.03 |
| Normal weight (18.5 to <25) | 245 (10.1%) | 3,011 (10.7%) | 0.02 | 241 (10.3%) | 246 (10.5%) | 0.01 |
| Overweight (25 to <30) | 504 (20.7%) | 4,154 (14.7%) | 0.17 | 488 (20.9%) | 476 (20.4%) | 0.01 |
| Obese – class 1 (30 to <35) | 419 (17.2%) | 3,330 (11.8%) | 0.17 | 410 (17.6%) | 456 (19.5%) | 0.05 |
| Obese – class 2 (35 to <40) | 383 (15.7%) | 1,552 (5.5%) | 0.42 | 357 (15.3%) | 379 (16.2%) | 0.03 |
| Obese – class 3 (≥40) | 443 (18.2%) | 1,208 (4.3%) | 0.62 | 399 (17.1%) | 380 (16.3%) | 0.02 |
| **Before Matching** |  |  |  | **After Matching** |  |  |
| **Clinical covariate** | **Bamlanivimab cohort**<br>**(n = 2,436)** | **Control cohort**<br>**(n = 28,230)** | **Standardized Difference** | **Bamlanivimab cohort**<br>**(n = 2,335)** | **Control cohort**<br>**(n = 2,335)** | **Standardized Difference** |
| Unknown | 435 (17.9%) | 14,859 (52.6%) | 0.71 | 434 (18.6%) | 388 (16.6%) | 0.05 |
| **Comorbidity** |  |  |  |  |  |  |
| Hypertension | 1,353 (55.5%) | 4,878 (17.3%) | 0.98 | 1,265 (54.2%) | 1,286 (55.1%) | 0.02 |
| Chronic Pulmonary Disease | 635 (26.1%) | 3,085 (10.9%) | 0.47 | 585 (25.1%) | 572 (24.5%) | 0.01 |
| Diabetes Mellitus w/o complications | 381 (15.6%) | 1,034 (3.7%) | 0.58 | 352 (15.1%) | 345 (14.8%) | 0.01 |
| Cancer (Local) | 375 (15.4%) | 1,062 (3.8%) | 0.56 | 342 (14.6%) | 337 (14.4%) | 0.01 |
| Peripheral Vascular Disease | 372 (15.3%) | 1,138 (4.0%) | 0.52 | 341 (14.6%) | 301 (12.9%) | 0.05 |
| Renal Disease | 369 (15.1%) | 1,027 (3.6%) | 0.56 | 339 (14.5%) | 280 (12.0%) | 0.07 |
| Diabetes Mellitus w/ complications | 295 (12.1%) | 694 (2.5%) | 0.55 | 267 (11.4%) | 213 (9.1%) | 0.08 |
| Liver disease (mild) | 255 (10.5%) | 911 (3.2%) | 0.38 | 230 (9.9%) | 204 (8.7%) | 0.04 |
| Congestive Heart Failure | 248 (10.2%) | 707 (2.5%) | 0.45 | 219 (9.4%) | 166 (7.1%) | 0.08 |
| Cerebrovascular Disease | 223 (9.2%) | 759 (2.7%) | 0.37 | 209 (9.0%) | 167 (7.2%) | 0.07 |
| Myocardial Infarction | 149 (6.1%) | 434 (1.5%) | 0.34 | 134 (5.7%) | 88 (3.8%) | 0.09 |
| Connective Tissue Disease | 144 (5.9%) | 406 (1.4%) | 0.34 | 133 (5.7%) | 123 (5.3%) | 0.02 |
| Cancer (Metastatic) | 73 (3.0%) | 263 (0.9%) | 0.2 | 67 (2.9%) | 70 (3.0%) | 0.01 |
| Peptic Ulcer Disease | 48 (2.0%) | 261 (0.9%) | 0.1 | 46 (2.0%) | 38 (1.6%) | 0.03 |
| Paraplegia / Hemiplegia | 23 (0.9%) | 107 (0.4%) | 0.09 | 23 (1.0%) | 21 (0.9%) | 0.01 |
| Dementia | 20 (0.8%) | 155 (0.5%) | 0.04 | 19 (0.8%) | 18 (0.8%) | 0 |
| Liver Disease<br>(Moderate/Severe) | 19 (0.8%) | 91<br>(0.3%) | 0.08 | 18 (0.8%) | 17 (0.7%) | 0 |
| HIV/AIDS | 6 (0.2%) | 17<br>(0.1%) | 0.07 | 5 (0.2%) | 3 (0.1%) | 0.02 |
| Immunosuppressant use | 165 (6.8%) | 354<br>(1.3%) | 0.98 | 143 (6.1%) | 116<br>(5.0%) | 0.05 |
| Time from PCR date to infusion (days) |  |  |  |  |  |  |
| Mean | 2.8 |  |  | 2.8 |  |  |
| Median | 2.0 |  |  | 2.0 |  |  |
| Range | (0, 10) |  |  | (0, 10) |  |  |

<sup>1</sup>Other race categories include: Samoan, Guamanian or Chamorro, Native Hawaiian/Pacific Islander, Other Pacific Islander, Unable to Provide, and Other.
IQR, interquartile range; HIV/AIDS, human immunodeficiency virus/acquired immunodeficiency syndrome

### Demographic and clinical covariates

To perform propensity matching described above, demographic and clinical covariates which could influence the likelihood of bamlanivimab administration were considered (**Table 1**). Demographic covariates considered included age, sex, race, and ethnicity. Race and ethnicity were determined based on patient entered responses to multiple choice questions with fixed categories and were considered in this study in order to control for social determinants of health and other potential confounding factors. Clinical covariates were derived from the Charlson Comorbidity Index and were identified for each patient on the basis of ICD-9 and ICD-10 codes recorded in the 5 years prior to the SARS-CoV-2 PCR testing date (**eTable 1**).

Other covariates considered during the propensity score matching included hypertension, BMI, immunosuppressive medication usage, and location of infusion. Hypertension status was determined using ICD-10 codes recorded in the 5 years prior to the PCR testing date (**eTable 1**). BMI was calculated using most recently recorded weight (between one year before and one week after COVID-19 diagnosis) and height (between age 18 and one week after COVID-19 diagnosis). Immunosuppressive medication usage was determined using medication orders active or completed in the year prior to the PCR testing date up to the end of the study period (**eTable 4**). This study included participants from four major sites: Scottsdale, Arizona; Jacksonville, Florida; Rochester, Minnesota; and other Mayo Clinic Health Systems sites. Location of infusion was incorporated into the covariate balancing analysis post-hoc. Due to the small number of sites, this variable was modelled as a fixed effect.^14^

### Outcomes

The clinical outcomes that were assessed between the bamlanivimab-treated and control cohorts at days 14, 21, and 28 after study enrollment were rates of hospitalization, ICU admission, and death, and number of hospital- and ICU-free days. Hospitalization rate was the primary outcome of interest. To determine the outcome for each cohort, only patients with sufficient follow-up data relative to the end study date (February 17, 2021) were considered. For example, to determine the 14-day outcomes for each cohort, only patients with enrollment dates on or prior to February 3, 2021 were included.

### Statistical analyses

Prior to the statistical analysis, missing values were imputed. Among all of the covariates, the only ones with missing data were race (0.8%), ethnicity (1.8%), and BMI (17.9%) (see **Table 1**). For covariates with missing data, the missing values were categorized as “Unknown”.

The effectiveness of covariate balancing between bamlanivimab-treated and control cohort was assessed using the standardized difference.^17,18^ To compare the rates of hospitalization, ICU admission, and death at the defined time points after study enrollment, the percentage of patients positive for each outcome relative to the total number of patients with follow-up in each cohort was calculated. For each of these outcome variables, a logistic regression model was fit using the treatment variable, controlling for all of the covariates considered during the propensity-matching step as potential residual confounding factors.^19^ For each outcome, the odds ratio (OR; bamlanivimab versus control) was computed along with the corresponding 95% confidence intervals (CI) and p-values. In addition, for each outcome, the absolute difference in days was reported, along with the 95% CI for this difference. The logistic regression models were implemented using the “statsmodels” package (v.0.10.0) in python.^20^ To test the robustness of study findings, post-hoc “negative outcome” and “intention-to-treat” sensitivity analyses were conducted, as described below.

To compare hospital-free and ICU-free days at the defined time points after study enrollment, the mean number of hospital-free and ICU-free days among patients with follow-up were calculated for each cohort, along with their 95% CI. The differences in means (95% CI) were calculated, and significance was assessed with a Mann-Whitney test. Because of the potential for type 1 error due to multiple comparisons, analysis of the secondary outcomes should be interpreted as exploratory. For each of the statistical tests, a two-sided p-value < 0.05 was considered statistically significant. Analysis was performed with the aid of the “scipy” package (v0.25.6) in Python.^22^

Hospitalization-free survival was also assessed at daily intervals with a Kaplan-Meier analysis and a corresponding log-rank test. Specifically, the proportion of patients in each cohort (among those with follow-up) who were not hospitalized on each day after study enrollment was compared (**eTable 3, Figure 2**). Survival analysis was performed using the “lifelines” package (v0.25.6) in Python.^23^

**Figure 2:**
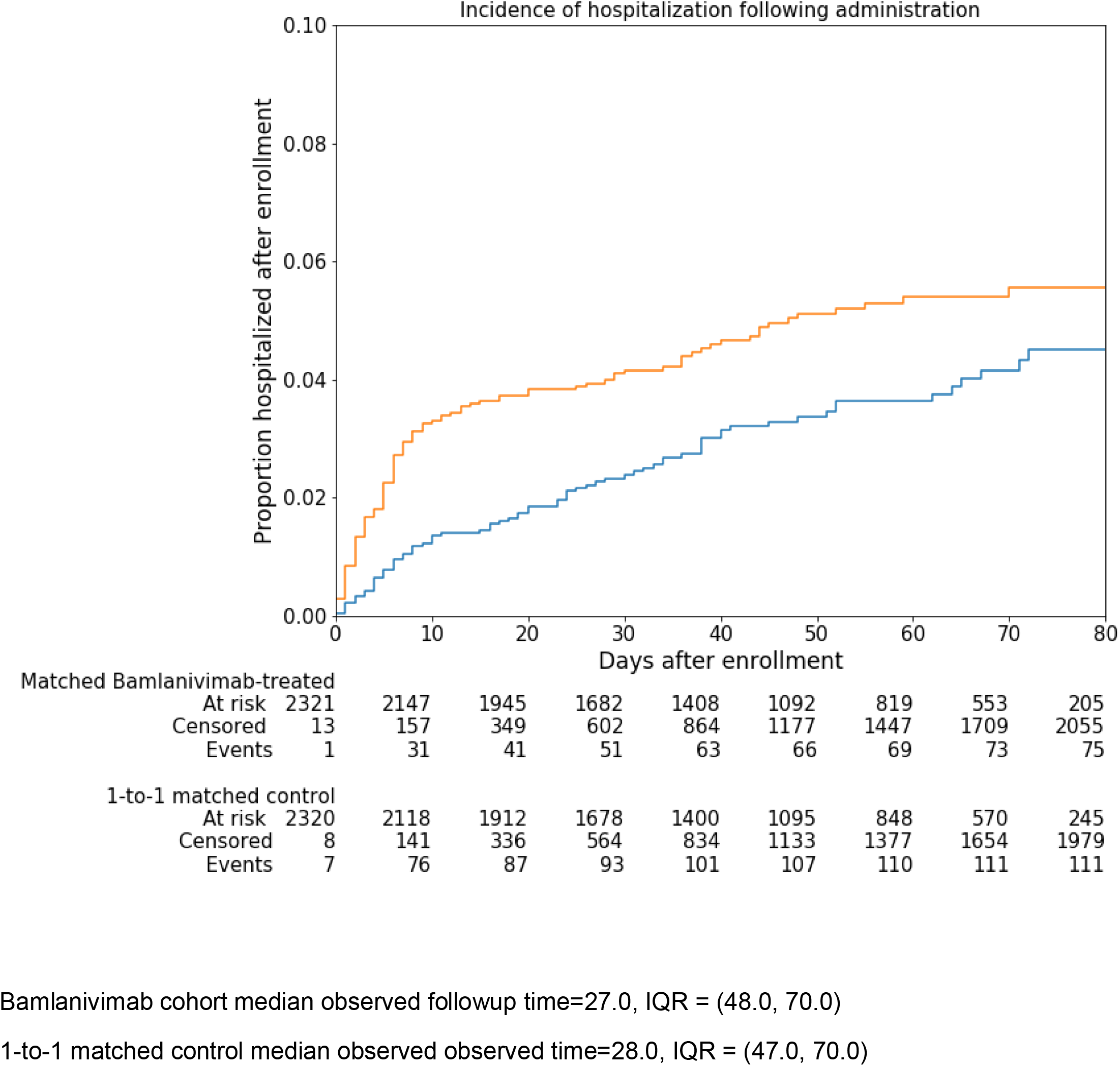
Cumulative Incidence of Hospitalization Over Time in Bamlanivimab-treated and Propensity-matched Untreated Control Population.

Post-hoc statistical tests were done to account the fact that the endpoints were measured at multiple timepoints. For each of the survival-type outcomes (hospital admission, ICU admission, and mortality), the p-values from 28-day capped log-rank tests between the propensity matched cohorts were reported. These tests are used to assess whether or not the hospital admission rates, ICU admission rates, and mortality rates are significantly different between the matched cohorts within the 28 day follow-up period. Log-rank tests were performed using the “lifelines” (v0.25.6) package in Python. For each of the numeric outcomes (number of hospital-free days and number of ICU-free days), we report p-values from two-way mixed ANOVA tests, using treatment (bamlanivimab or control) as the between-subjects factor and time point (14, 21, or 28 days) as the within-subjects factor.^24^ Mixed ANOVA tests were performed using the “pingouin” (v0.3.11) package in Python, with the Greenhouse-Geisser correction applied afterwards to adjust for violations of the sphericity assumption.

### Sensitivity analysis

Two post-hoc sensitivity analyses were performed to test the robustness of the findings. First, in order to test the sensitivity of the study findings to potential unobserved confounding variables, the statistical analysis was repeated on a “negative outcome” suspected to be unrelated to COVID-19 disease or treatment. For this post-hoc sensitivity analysis, cancer screening was considered as the negative outcome. In particular, the negative outcome was defined as +1 for patients with an ICD-10 diagnostic code for cancer screening (Z12.*) 15 to 42 days following their PCR diagnosis date, and 0 otherwise. Patients with PCR diagnosis dates after February 8, 2021 were excluded from this sensitivity analysis due to lack of 42-day follow-up data.

Second, an “intention-to-treat” sensitivity analysis was conducted to compare the outcomes between all patients who received an order for bamlanivimab versus the control group. In this analysis, all patients with cancelled orders for bamlanivimab was considered in addition to the bamlanivimab-treated patients, subjected to the same inclusion criteria. For each of the patients with cancelled bamlanivimab orders, the relative infusion dates were randomly sampled from the distribution of relative infusion dates for the actual bamlanivimab-treated cohort, ensuring that the infusion dates were within the study period. A 1:1 propensity matching was performed between the bamlanivimab intended-to-treat cohort and the control cohort, following the same procedure as in the primary analysis. The rates of hospitalization, ICU admission, and mortality for the matched cohorts were compared for the 14, 21, and 28-day time horizons.

## Results

### Patient population

Of the 33,446 adult patients with positive SARS-CoV-2 PCR test, the participant selection algorithm (**Figure 1**) resulted into two cohorts that were balanced for relevant demographic and clinical covariates: (1) bamlanivimab-treated patients (n = 2,335), and (2) control patients (n = 2,335) who did not receive monoclonal antibody (**Table 1)**. Appropriate matches could not be found for 101 bamlanivimab-treated patients which led to a decrease in size of our bamlanivimab cohort from 2,436 to 2,335. Unmatched data is shown in **eTable 2** and the effect of propensity score matching on these variables is shown in **eTable 3**. All covariates showed standardized differences < 0.1 confirming that the cohorts were reasonably balanced for reliable downstream comparisons (**Table 1**). The success of balancing was also confirmed by comparing the age distribution (**eFigure 3**) and the prevalence of each categorical covariate (**eFigure 4**) in the two cohorts before and after propensity matching. Distribution of test results across the two cohorts is shown in **eFigure 5**.

The mean time from PCR date to bamlanivimab infusion was 2.79 days (median, 2 days) (**eFigure 4**).The most common comorbidities were hypertension (54.2%), diabetes mellitus (26.5%), chronic lung disease (25.1%), renal disease (14.5%), malignancy (16.6%), peripheral vascular disease (14.6%), liver disease (9.6%), congestive heart failure (9.0%), and immunosuppressive drug use (6.1%).

### Primary Outcome

#### All-cause hospitalization

All-cause hospitalization rates were significantly lower in the bamlanivimab group than the propensity-matched cohort at days 14 (1.5% vs 3.5% (difference, 2.1%; 95% confidence interval [CI]: 1.2%-3.0%);Odds Ratio [OR], 0.38; 95% CI: 0.25-0.59), 21 (1.9% vs 3.9%(difference, 2.0%; 95% CI: 0.91%-3.0%); OR, 0.46; 95% CI: 0.30-0.70), and 28 (2.5% vs 3.9% (difference 1.5%; 95% CI: 0.33%-2.6%); OR, 0.61; 95% CI: 0.41-0.91) (**Table 2**). Bamlanivimab-treated patients had significantly more hospitalization-free days at all time points, compared to the propensity-matched cohort (**Table 3**). Kaplan-Meier survival analysis showed significant separation in rate of hospitalization-free survival between the bamlanivimab-treated and propensity-matched control (log-rank test p-value 0.01; **Figure 2**).

**Table 2:** Hospitalizations, intensive care unit admissions, and mortality for bamlanivimab-treated and untreated control cohort.

| Outcome | Bamlanivimab (n = 2335) | Control (n = 2335) | Risk difference (95% CI) | Odds ratio (95% CI) | Odds Ratio p-value<sup>†</sup> | Log-rank p-value |
| --- | --- | --- | --- | --- | --- | --- |
| **Number of patients with follow up data** |  |  |  |  |  |  |
| **14 day** | **2126** | 2145 |  |  |  |  |
| **21 day** | 1983 | 1989 |  |  |  |  |
| **28 day** | 1789 | 1832 |  |  |  |  |
| **Hospitalizations** |  |  |  |  |  |  |
| **14 day** | 31 (1.5%) | 76 (3.5%) | 2.1% (1.2%, 3%) | 0.38 (0.25, 0.59) | <0.001 | <0.001 |
| **21 day** | 38 (1.9%) | 77 (3.9%) | 2% (0.91%, 3%) | 0.46 (0.30, 0.70) | <0.001 |  |
| **28 day** | 44 (2.5%) | 72 (3.9%) | 1.5% (0.33%, 2.6%) | 0.61 (0.41, 0.91) | 0.02 |  |
| **Intensive Care Unit admissions** |  |  |  |  |  |  |
| **14 day** | 3 (0.14%) | 22 (1%) | 0.88% (0.43%, 1.3%) | 0.12 (0.03, 0.43) | 0.001 | 0.06 |
| **21 day** | 5 (0.25%) | 20 (1%) | 0.75% (0.26%, 1.2%) | 0.24 (0.08, 0.66) | 0.006 |  |
| **28 day** | 10 (0.56%) | 20 (1.1%) | 0.53% (-0.055%, 1.1%) | 0.52 (0.23, 1.16) | 0.11 |  |
| **Mortality** |  |  |  |  |  |  |
| **14 day** | 0 (0%) | 7 (0.33%) | 0.33% (0.085%, 0.57%) | N/A | 1 | 0.06 |
| **21 day** | 1 (0.05%) | 8 (0.4%) | 0.35% (0.057%, 0.65%) | 0.08 (0.01, 0.82) | 0.03 |  |
| **28 day** | 2 (0.11%) | 8 (0.44%) | 0.32% (-0.014%, 0.66%) | 0.01 (0.00, 0.37) | 0.01 |  |

<sup>†</sup>P-values for the null hypothesis that the odds ratio is 1.

**Table 3:** Hospitalization-free days and intensive care unit-free days for bamlanivimab-treated vs. untreated control group.

| Outcome | Bamlanivimab<br>(n = 2335) |  | Control<br>(n = 2335) |  | Absolute<br>difference<br>(95% CI) | Mann-Whitney p-value | Two-way<br>mixed<br>ANOVA p-value |
| --- | --- | --- | --- | --- | --- | --- | --- |
|  | Mean<br>(SD) | Median<br>(IQR) | Mean<br>(SD) | Median<br>(IQR) |  |  |  |
| Hospital-free days |  |  |  |  |  |  |  |
| 14 day | 13.9 (0.6) | 14 [14, 14] | 13.8 (1.3) | 14 [14, 14] | 0.1 (0.1, 0.2) | <0.001 | 0.01 |
| 21 day | 20.9 (0.8) | 21 [21, 21] | 20.7 (1.8) | 21 [21, 21] | 0.2 (0.1, 0.3) | <0.001 |  |
| 28 day | 27.9 (0.9) | 28 [28,28] | 27.7 (2.3) | 28 [28,28] | 0.2 (0.1, 0.3) | 0.01 |  |
| Intensive Care Unit-free days |  |  |  |  |  |  |  |
| 14 day | 14.0 (0.1) | 14 [14, 14] | 13.9 (0.6) | 14 [14, 14] | 0.1 (0.0, 0.1) | <0.001 | 0.005 |
| 21 day | 21.0 (0.1) | 21 [21, 21] | 20.9 (1.1) | 21 [21, 21] | 0.1 (0.0, 0.1) | 0.003 |  |
| 28 day | 28.0 (0.2) | 28 [28,28] | 27.9 (1.6) | 28 [28,28] | 0.1 (0.1, 0.2) | 0.07 |  |

SD, standard deviation; IQR, interquartile range; 95%CI, 95% confidence intervals

### Secondary Outcomes

#### Intensive care unit admissions

All-cause ICU admission rates were lower in bamlanivimab-treated compared to the propensity-matched cohort at days 14 (0.14% vs 1%, (difference 0.88%; 95% CI: 0.43%-1.3%); OR, 0.12; 95% CI: 0.03-0.43), 21 (0.25% vs 1%, (difference 0.75%; 95% CI: 0.26%-1.2%); OR: 0.24; 95% CI: 0.08-0.66) and 28 (0.56% vs 1.1%, (difference 0.53%; 95% CI: -0.06%-1.1%); OR: 0.52; 95% CI: 0.23-1.16) (**Table 2**). Bamlanivimab-treated patients had significantly more ICU-free days, at days 14 and 21, compared to the propensity-matched cohort (**Table 3**). Ventilator days were similar between the two cohorts with mechanical ventilation required in one of 10 ICU-admitted patients in the bamlanivimab group and two of 19 ICU-admitted patients in the control group.

#### Survival

Patients treated with bamlanivimab had lower all-cause mortality compared to the propensity-matched cohort at days 14 (0% vs 0.33%, (difference 0.33%; 95% CI: 0.09%-1.1%)), 21 (0.05% vs 0.4%, (difference 0.35%, 95% CI: 0.06%-0.65%); OR,0.08; 95% CI: 0.01-0.82) and 28 (0.11% vs 0.44%, difference 0.32%, 95% CI: 0.01%-0.66%); OR, 0.01; 95% CI: 0.00-0.37). Only two bamlanivimab-treated patients (among 1789 patients with at least 28 days of follow up) died, on days 20 and 25, of causes unrelated to COVID-19. In the untreated cohort, 7 of 8 deaths were attributable to COVID-19.

### Sensitivity analyses

#### Falsification Outcome

**eTable 5** shows the results comparing the negative outcomes in the original and propensity-matched cohorts. Prior to matching, the difference in cancer screening rates between the treated and untreated cohorts was statistically significant (Fisher exact test p-value: 0.002), but after matching, the difference in rates was not statistically significant (Fisher exact test p-value: 0.67). In addition, in both cases the difference in rates was not statistically significant after controlling for residual confounding factors via logistic regression. This demonstrated that the matching procedure was effective in controlling for potential confounding factors which may lead the treated or untreated cohorts to be enriched for non-COVID-19 related endpoints.

#### Intention to Treat Sensitivity Analysis

There were 183 additional patients with cancelled orders for bamlanivimab included in the intended-to-treat analysis. After 1:1 propensity matching, there were 2,509 patients in the bamlanivimab intended-to-treat cohort, and 2,509 patients in the control cohort. **eTable 6** showed the results comparing the hospitalization, ICU admission, and mortality outcomes of these cohorts. The 14-day and 21-day outcomes for hospitalization and ICU admission remained statistically significant for this comparison. These results suggested that it is unlikely that treatment cancellation bias has strongly influenced the study findings.

### Adverse Events

Adverse events were reported in 19 patients with fever and chills (n=6), nausea and vomiting (n=5), and lightheadedness (n=3) being most common. Rash, chest pain, confusion, weakness (2 each) and diarrhea, headache, cough, facial swelling, and dyspnea (1 each) were also observed. No one had anaphylaxis. All adverse events were mild and did not require hospitalization.

### Impact of Therapy

Based on this study, it is estimated that, in the first 28 days of follow up of 1789 patients, there were 358 hospital days, 179 ICU days, and 6 lives saved.

## Discussion

This retrospective study shows that bamlanivimab monotherapy was associated with a statistically significant decrease in the rate of all-cause hospitalization at 28 days after infusion, with greater effects demonstrated at 14 and 21 days.^25^

Among high-risk patients 65 years and older and those with BMI >35 who participated in the phase 2 randomized clinical trial, the rate of hospitalization and medically-attended visit was 4% among bamlanivimab-treated patients compared to 15% among those who received placebo.^6^ Our reported rates in this study are numerically lower, especially among the untreated propensity-matched control group.^6^ This difference could have been due to the different time point of enrollment, as the current cohort represents a contemporary population when the care of COVID-19 patients has improved. The lower rate of hospitalization in the untreated cohort lessened the anticipated magnitude of impact of bamlanivimab therapy. Nonetheless, this study observed that bamlanivimab was significantly associated with reduction in all-cause hospitalizations, by 57% at day 14 and 31% by day 28.

This study suggests that bamlanivimab-treated patients were less likely to progress to critical illness, supported by the lower rates of intensive care unit admission and mortality, notable in a cohort that included only the high-risk population. ^26,27^ By virtue of the strict FDA EUA criteria, 100% of the bamlanivimab-treated population had at least one risk factor for severe COVID-19.^6^ As prior studies have suggested that patients with medical comorbidities are at higher risk of severe and critical illness, early treatment with bamlanivimab may have mitigated this progression.^28,29^

Due to the evolution of SARS-CoV-2 variants, bamlanivimab is no longer authorized for use as monotherapy. Despite this, the results of this study which showed the real-world efficacy of bamlanivimab monotherapy in the prevention of hospital admission and other serious clinical outcomes, provides proof that treatment with neutralizing antibodies is an effective strategy to mitigate the current COVID-19 pandemic. The observations in this study could serve as a model that may be translatable to the use of other monoclonal antibody combination therapies which are still available.

## Limitations

This study has several limitations. First, this was an observational cohort study, and precludes the causal inference that can result from a randomized clinical trial. However, performing a randomized trial was not feasible due to the ethical implications of withholding a drug authorized for emergency use in the treatment of high-risk patients. Propensity score matching was performed in an attempt to reduce confounding bias. Furthermore, two sensitivity analyses demonstrated that the matching procedure was effective in controlling for potential confounding variables. Second, this study had a retrospective design, and may not have captured all the clinical outcomes of patients who may have received subsequent care in other institutions. This limitation is mitigated by the extensive outpatient remote monitoring and follow up program.^30^ Also, only patients with documented follow up were included in the analysis of outcomes at days 14, 21 and 28. Third, this study focused on bamlanivimab monotherapy and did not include patients who received casirivimab and imdevimab or bamlanivimab and etesevimab combination therapy. The clinical outcomes reported here therefore only apply to one specific monoclonal antibody, for which the EUA has been revoked secondary to resistance patterns of emerging SARS-CoV-2 variants in the community.^5^ Fourth, the study population was predominantly White, and further studies will need to be performed to validate the findings in other populations. Fifth, the outcomes data was derived from a single multi-site healthcare system which proactively screened and consented all eligible patients leading to rapid infusion of monoclonal antibody, and thus, the results may not be generalizable to other systems with different practices. Sixth, despite the large patient population and the statistical significance, the magnitude of some findings is small. In particular, the difference in ICU-free and hospitalization-free days are small when considered at the patient level. However, this small difference can be magnified when considered at the larger population level.

## Conclusions

Among high-risk patients with mild to moderate COVID-19, treatment with bamlanivimab compared with usual care was associated with statistically significant lower rate of hospitalization at 28 days. There were also significant reductions observed in the rates of ICU admissions and mortality. While bamlanivimab monotherapy is no longer authorized, the observations in this study provides proof that treatment with neutralizing antibodies are effective clinically in reducing hospitalization, ICU admission and mortality in patients with mild to moderate COVID-19. We suggest a similar real-world study to assess the clinical outcomes of currently authorized combination anti-spike monoclonal antibody therapies.

## Data Availability

Data referred to in the manuscript is maintained by the authors and available upon request

## Author Contributions

<u>Concept and Design:</u> Ganesh, Razonable, Pawlowski

<u>Acquisition, Analysis, or Interpretation of Data:</u> Pawlowski, Lenehan, Puranik, Venkatakrishnan, O’Horo, Ganesh, Razonable, Badley

<u>Drafting of the Manuscript:</u> Ganesh, Razonable, Pawlowski

<u>Critical Revision of the Manuscript:</u> all authors

<u>Statistical Analysis:</u> Pawlowski, Lenehan, Puranik, Venkatakrishnan, O’Horo, Ganesh, Razonable

<u>Administrative, Technical, or Material Support:</u> Bell, Larsen, Destro Borgen, Badley, Moehnke,

<u>Supervision:</u> Orenstein, Speicher, Tulledge-Scheitel, Wilker, Hanson, Bierle, Ganesh, Razonable

## Acknowledgments

Additional members of the MATRx team are listed in the **eSupplement**. They are all affiliated with the Mayo Clinic and did not receive compensation for their work in establishing the Mayo Clinic Monoclonal Antibody Infusion Program. Mayo Clinic as the funding entity did not have any role in the design and conduct of the study; collection, management, analysis, and interpretation of the data; preparation, review, or approval of the manuscript; and decision to submit the manuscript for publication. Mayo Clinic did not retain the right to veto publication or dictate which journal the paper should be submitted to.

