## supplemental tables and figures for "Association of Intravenous Bamlanivimab Use with Reduced Hospitalization, Intensive Care Unit Admission, and Mortality in Patients with Mild to Moderate COVID-19"

**TABLE OF CONTENTS:**

| Title | Page |
| --- | --- |
| eFigure 1 | 2 |
| eFigure 2 | 3 |
| eFigure 3 | 4 |
| eFigure 4 | 5 |
| eFigure 5 | 6 |
| eTable 1 | 7 |
| eTable 2 | 11 |
| eTable 3 | 12 |
| eTable 4 | 13 |
| eTable 5 | 14 |
| eTable 6 | 15 |
| eFigure 6 | 16 |
| MATRx team members | 17 |
| References | 18 |

**eFigure 1: Infusion dates among bamlanivimab patients (n=2,436) relative to first positive PCR**

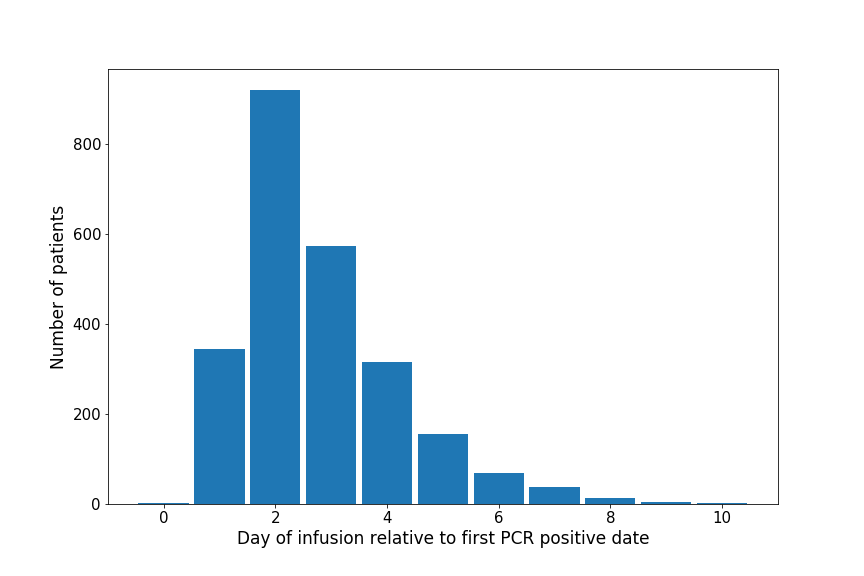

Mean time to infusion was 2.79 days (median 2.0 days)

**eFigure 2: Distribution of relative enrollment date of matched control relative to bamlanivimab infusion date (n=2355).** In the matching procedure, the matched control for a bamlanivimab-treated patient was restricted to those patients with study enrollment date within 2 weeks (and preferably 1 week) of the bamlanivimab-treated patient -- equivalently, the first SARS-CoV-2 PCR positive date of the matched control patient must be within 2 weeks of the first PCR positive date of the bamlanivimab-treated patient. *Relative enrollment date* is defined as the difference between the matched control enrollment and the bamlinivimab-treated patient’s infusion: so if the matched control is enrolled 6 days prior to the bamlinivimab-treated patient’s infusion date, this counts as a relative enrollment date of -6. 2,264 of the 2,355 matched control patients have relative enrollment between -7 and 7. 

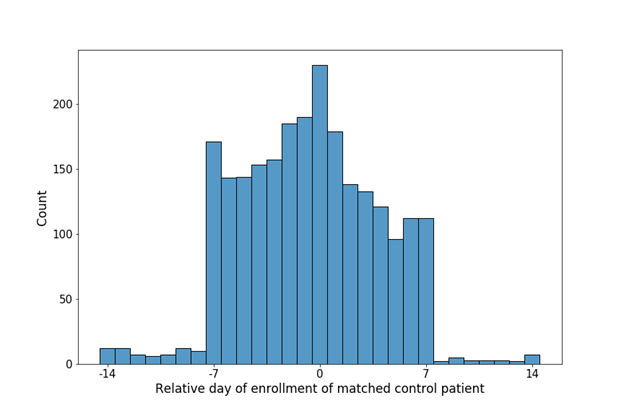

**eFigure 3: Age balance before and after propensity score matching.** (A) before matching, (B) after matching. Blue denotes bamlanivimab-treated, orange denotes control patients.

A. **Before matching**

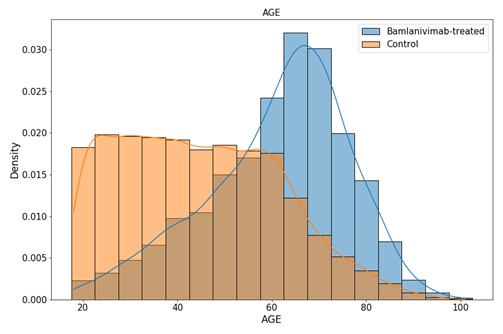

B. **After matching**

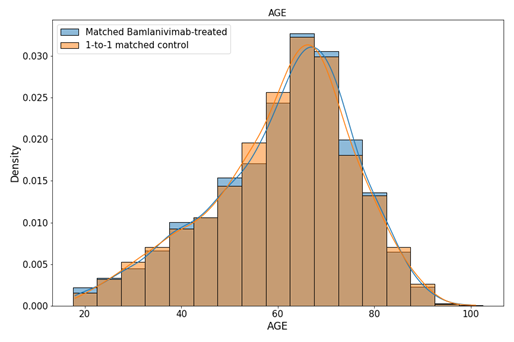

**eFigure 4: Binary covariate prevalences before and after propensity score matching.** (A) before matching and (B) after matching. Blue denotes bamlanivimab-treated, orange denotes control patients.

A. **Before matching**

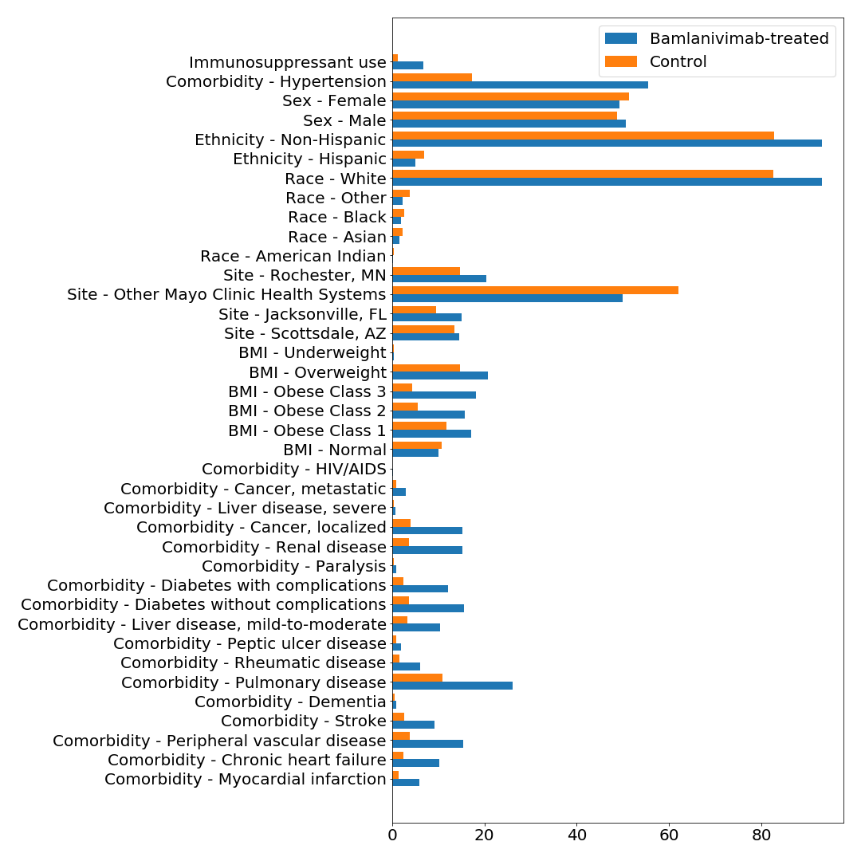

B. **After matching**

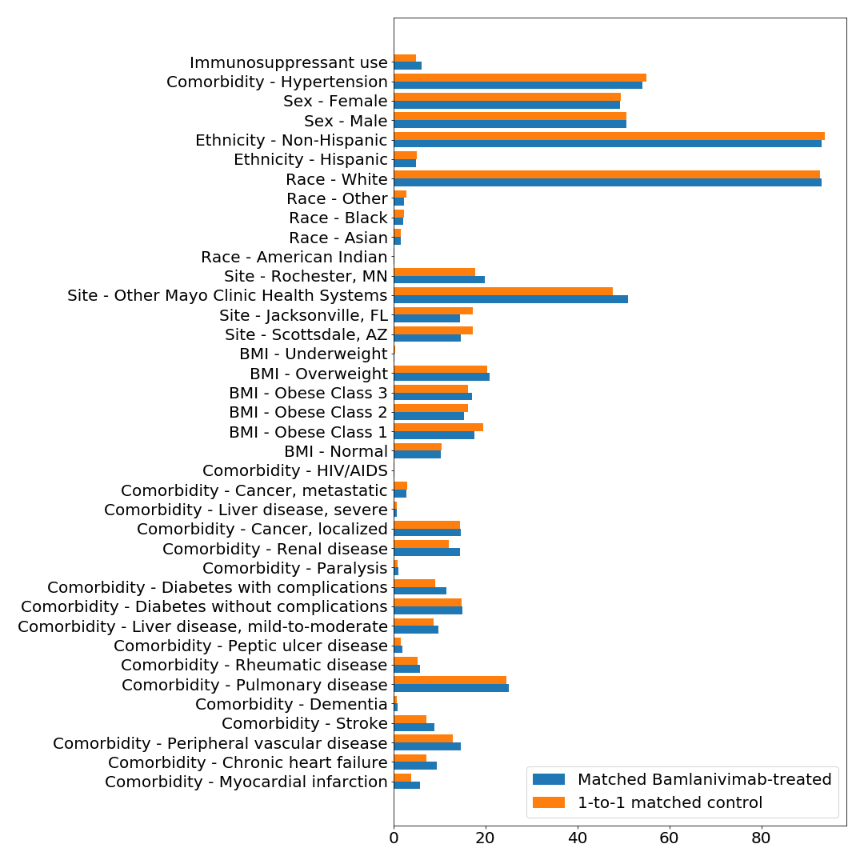

**eFigure 5: Distribution of PCR diagnosis dates for propensity matched cohorts.**

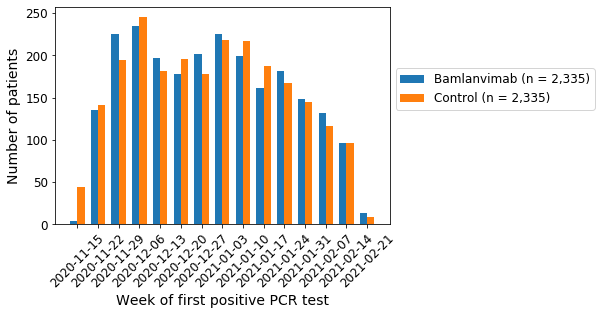

**eTable 1: ICD diagnostic codes for Charlson comorbidities and hypertension.**

| **Comorbidity** | **ICD-10 code** | **Description** |
| --- | --- | --- |
| LiverMild | B18 | Chronic viral hepatitis |
| HIV | B20 | Human immunodeficiency virus [HIV] disease |
| Cancer | C00 | Malignant neoplasm of lip |
| Cancer | C01 | Malignant neoplasm of base of tongue |
| Cancer | C02 | Malignant neoplasm of other and unspecified parts of tongue  Malignant neoplasm of other and unspecified parts of tongue |
| Cancer | C03 | Malignant neoplasm of gum |
| Cancer | C04 | Malignant neoplasm of floor of mouth |
| Cancer | C05 | Malignant neoplasm of palate |
| Cancer | C06 | Malignant neoplasm of other and unspecified parts of mouth   Malignant neoplasm of other and unspecified parts of mouth |
| Cancer | C07 | Malignant neoplasm of parotid gland |
| Cancer | C08 | Malignant neoplasm of other and unsp major salivary glands   Malignant neoplasm of other and unspecified major salivary glands |
| Cancer | C09 | Malignant neoplasm of tonsil |
| Cancer | C10 | Malignant neoplasm of oropharynx |
| Cancer | C11 | Malignant neoplasm of nasopharynx |
| Cancer | C12 | Malignant neoplasm of pyriform sinus |
| Cancer | C13 | Malignant neoplasm of hypopharynx |
| Cancer | C14 | Malig neoplasm of sites in the lip, oral cavity and pharynx  Malignant neoplasm of other and ill-defined sites in the lip, oral cavity and pharynx |
| Cancer | C15 | Malignant neoplasm of esophagus |
| Cancer | C16 | Malignant neoplasm of stomach |
| Cancer | C17 | Malignant neoplasm of small intestine |
| Cancer | C18 | Malignant neoplasm of colon |
| Cancer | C19 | Malignant neoplasm of rectosigmoid junction |
| Cancer | C20 | Malignant neoplasm of rectum |
| Cancer | C21 | Malignant neoplasm of anus and anal canal |
| Cancer | C22 | Malignant neoplasm of liver and intrahepatic bile ducts   Malignant neoplasm of liver and intrahepatic bile ducts |
| Cancer | C23 | Malignant neoplasm of gallbladder |
| Cancer | C24 | Malignant neoplasm of other and unsp parts of biliary tract  Malignant neoplasm of other and unspecified parts of biliary tract |
| Cancer | C25 | Malignant neoplasm of pancreas |
| Cancer | C26 | Malignant neoplasm of other and ill-defined digestive organs Malignant neoplasm of other and ill-defined digestive organs |
| Cancer | C30 | Malignant neoplasm of nasal cavity and middle ear |
| Cancer | C31 | Malignant neoplasm of accessory sinuses |
| Cancer | C32 | Malignant neoplasm of larynx |
| Cancer | C33 | Malignant neoplasm of trachea |
| Cancer | C34 | Malignant neoplasm of bronchus and lung |
| Cancer | C37 | Malignant neoplasm of thymus |
| Cancer | C38 | Malignant neoplasm of heart, mediastinum and pleura |
| Cancer | C39 | Malig neoplm of sites in the resp sys and intrathorac organs Malignant neoplasm of other and ill-defined sites in the respiratory system and intrathoracic organs |
| Cancer | C40 | Malignant neoplasm of bone and articular cartilage of limbs  Malignant neoplasm of bone and articular cartilage of limbs |
| Cancer | C41 | Malignant neoplasm of bone/artic cartl of and unsp sites  Malignant neoplasm of bone and articular cartilage of other and unspecified sites |
| Cancer | C43 | Malignant melanoma of skin |
| Cancer | C45 | Mesothelioma |
| **Comorbidity** | **ICD-10 code** | **Description** |
| Cancer | C46 | Kaposi's sarcoma |
| Cancer | C47 | Malignant neoplasm of prph nerves and autonomic nervous sys  Malignant neoplasm of peripheral nerves and autonomic nervous system |
| Cancer | C48 | Malignant neoplasm of retroperitoneum and peritoneum |
| Cancer | C49 | Malignant neoplasm of other connective and soft tissue |
| Cancer | C50 | Malignant neoplasm of breast |
| Cancer | C51 | Malignant neoplasm of vulva |
| Cancer | C52 | Malignant neoplasm of vagina |
| Cancer | C53 | Malignant neoplasm of cervix uteri |
| Cancer | C54 | Malignant neoplasm of corpus uteri |
| Cancer | C55 | Malignant neoplasm of uterus, part unspecified |
| Cancer | C56 | Malignant neoplasm of ovary |
| Cancer | C57 | Malignant neoplasm of other and unsp female genital organs   Malignant neoplasm of other and unspecified female genital organs |
| Cancer | C58 | Malignant neoplasm of placenta |
| Cancer | C60 | Malignant neoplasm of penis |
| Cancer | C61 | Malignant neoplasm of prostate |
| Cancer | C62 | Malignant neoplasm of testis |
| Cancer | C63 | Malignant neoplasm of other and unsp male genital organs  Malignant neoplasm of other and unspecified male genital organs |
| Cancer | C64 | Malignant neoplasm of kidney, except renal pelvis |
| Cancer | C65 | Malignant neoplasm of renal pelvis |
| Cancer | C66 | Malignant neoplasm of ureter |
| Cancer | C67 | Malignant neoplasm of bladder |
| Cancer | C68 | Malignant neoplasm of other and unspecified urinary organs   Malignant neoplasm of other and unspecified urinary organs |
| Cancer | C69 | Malignant neoplasm of eye and adnexa |
| Cancer | C70 | Malignant neoplasm of meninges |
| Cancer | C71 | Malignant neoplasm of brain |
| Cancer | C72 | Malig neoplm of spinal cord, cranial nerves and oth prt cnsl Malignant neoplasm of spinal cord, cranial nerves and other parts of central nervous system |
| Cancer | C73 | Malignant neoplasm of thyroid gland |
| Cancer | C75 | Malignant neoplasm of endo glands and related structures  Malignant neoplasm of other endocrine glands and related structures |
| Cancer | C76 | Malignant neoplasm of other and ill-defined sites |
| Mets | C77 | Secondary and unspecified malignant neoplasm of lymph nodes  Secondary and unspecified malignant neoplasm of lymph nodes |
| Mets | C78 | Secondary malignant neoplasm of resp and digestive organs Secondary malignant neoplasm of respiratory and digestive organs |
| Mets | C79 | Secondary malignant neoplasm of other and unspecified sites  Secondary malignant neoplasm of other and unspecified sites |
| Cancer | C7A | Malignant neuroendocrine tumors |
| Cancer | C7B | Secondary neuroendocrine tumors |
| Mets | C80 | Malignant neoplasm without specification of site |
| Cancer | C81 | Hodgkin lymphoma |
| Cancer | C82 | Follicular lymphoma |
| Cancer | C83 | Non-follicular lymphoma |
| Cancer | C84 | Mature T/NK-cell lymphomas |
| Cancer | C85 | Oth and unspecified types of non-Hodgkin lymphoma |
| Cancer | C88 | Malig immunoproliferative dis and certain oth B-cell lymph   Malignant immunoproliferative diseases and certain other B-cell lymphomas |
| Cancer | C90 | Multiple myeloma and malignant plasma cell neoplasms |
| **Comorbidity** | **ICD-10 code** | **Description** |
| Cancer | C91 | Lymphoid leukemia |
| Cancer | C92 | Myeloid leukemia |
| Cancer | C93 | Monocytic leukemia |
| Cancer | C94 | Other leukemias of specified cell type |
| Cancer | C95 | Leukemia of unspecified cell type |
| Cancer | C96 | Oth & unsp malig neoplm of lymphoid, hematpoetc and rel tiss Other and unspecified malignant neoplasms of lymphoid, hematopoietic and related tissue |
| Dementia | F01 | Vascular dementia |
| Dementia | F02 | Dementia in other diseases classified elsewhere |
| Dementia | F03 | Unspecified dementia |
| Dementia | G30 | Alzheimer's disease |
| Stroke | G45 | Transient cerebral ischemic attacks and related syndromes Transient cerebral ischemic attacks and related syndromes |
| Stroke | G46 | Vascular syndromes of brain in cerebrovascular diseases   Vascular syndromes of brain in cerebrovascular diseases |
| Paralysis | G81 | Hemiplegia and hemiparesis |
| Paralysis | G82 | Paraplegia (paraparesis) and quadriplegia (quadriparesis) Paraplegia (paraparesis) and quadriplegia (quadriparesis) |
| MI | I21 | Acute myocardial infarction |
| MI | I22 | Subsequent STEMI & NSTEMI mocard infrc |
| CHF | I43 | Cardiomyopathy in diseases classified elsewhere |
| CHF | I50 | Heart failure |
| Stroke | I61 | Nontraumatic intracerebral hemorrhage |
| Stroke | I62 | Other and unspecified nontraumatic intracranial hemorrhage   Other and unspecified nontraumatic intracranial hemorrhage |
| Stroke | I63 | Cerebral infarction |
| Stroke | I65 | Occls and stenosis of precerb art, not rslt in cereb infrc   Occlusion and stenosis of precerebral arteries, not resulting in cerebral infarction |
| Stroke | I66 | Occls and stenosis of cereb art, not rslt in cerebral infrc  Occlusion and stenosis of cerebral arteries, not resulting in cerebral infarction |
| Stroke | I67 | Other cerebrovascular diseases |
| Stroke | I68 | Cerebrovascular disorders in diseases classified elsewhere   Cerebrovascular disorders in diseases classified elsewhere |
| Stroke | I69 | Sequelae of cerebrovascular disease |
| PVD | I70 | Atherosclerosis |
| PVD | I71 | Aortic aneurysm and dissection |
| Pulmonary | J40 | Bronchitis, not specified as acute or chronic |
| Pulmonary | J41 | Simple and mucopurulent chronic bronchitis |
| Pulmonary | J42 | Unspecified chronic bronchitis |
| Pulmonary | J43 | Emphysema |
| Pulmonary | J44 | Other chronic obstructive pulmonary disease |
| Pulmonary | J45 | Asthma |
| Pulmonary | J47 | Bronchiectasis |
| Pulmonary | J60 | Coalworker's pneumoconiosis |
| Pulmonary | J61 | Pneumoconiosis due to asbestos and other mineral fibers   Pneumoconiosis due to asbestos and other mineral fibers |
| Pulmonary | J62 | Pneumoconiosis due to dust containing silica |
| Pulmonary | J63 | Pneumoconiosis due to other inorganic dusts |
| Pulmonary | J64 | Unspecified pneumoconiosis |
| Pulmonary | J65 | Pneumoconiosis associated with tuberculosis |
| Pulmonary | J66 | Airway disease due to specific organic dust |
| Pulmonary | J67 | Hypersensitivity pneumonitis due to organic dust |
| **Comorbidity** | **ICD-10 code** | **Description** |
| PUD | K25 | Gastric ulcer |
| PUD | K26 | Duodenal ulcer |
| PUD | K27 | Peptic ulcer, site unspecified |
| PUD | K28 | Gastrojejunal ulcer |
| LiverMild | K73 | Chronic hepatitis, not elsewhere classified |
| LiverMild | K74 | Fibrosis and cirrhosis of liver |
| Rheumatic | M05 | Rheumatoid arthritis with rheumatoid factor |
| Rheumatic | M06 | Other rheumatoid arthritis |
| Rheumatic | M32 | Systemic lupus erythematosus (SLE) |
| Rheumatic | M33 | Dermatopolymyositis |
| Rheumatic | M34 | Systemic sclerosis [scleroderma] |
| Renal | N19 | Unspecified kidney failure |
| Hypertension | I10 | Essential (primary) hypertension |
| Hypertension | I11 | Hypertensive heart disease |
| Hypertension | I12 | Hypertensive chronic kidney disease |
| Hypertension | I13 | Hypertensive heart and chronic kidney disease |
| Hypertension | I15 | Secondary hypertension |

For each comorbidity, we provide the ICD-10 diagnostic codes which were used to determine the presence of comorbidity in the bamlanivimab and control populations in the past 5 years relative to the first positive PCR testing date.  Only higher level codes are shown.

**eTable 2: Clinical characteristics of *unmatched* Bamlanvimab and control cohorts.**

| **Clinical covariate** | **Bamlanivimab cohort**  (2,436 patients) | **Control cohort**  (28,230 patients) |
| --- | --- | --- |
| Age (years)  < 65  65-75  > 75 | 1,274 (52.3%)  734 (30.1%)  428 (17.6%) | 24,566 (87.0%)  2,347 (8.3%)  1,317 (4.7%) |
| Sex  Female  Male | 1201 (49.3%)  1235 (50.7%) | 14,476 (51.3%)  13,746 (48.7%) |
| Race  American Indian  Asian  Black / African American  Other  White / Caucasian  Unknown | 4 (0.2%)  37 (1.5%)  49 (2.0%)  2,270 (93.2%)  56 (2.3%)  20 (0.8%) | 115 (0.4%)  633 (2.2%)  761 (2.7%)  23,302 (82.5%)  1,091 (3.9%)  2,328 (8.2%) |
| Ethnicity  Hispanic  Non-hispanic  Unknown | 121 (5.0%)  2,270 (93.2%)  45 (1.8%) | 1,969 (7.0%)  23,359 (82.7%)  2,902 (10.3%) |
| BMI (kg/m^2^)  Underweight (< 18.5)  Normal weight (18.5 to 25)  Overweight (25 to 30)  Obese - class 1 (30 to 35)  Obese - class 2 (35 to 40)  Obese - class 3 (≥ 40)  Unknown | 7 (0.3%)  245 (10.1%)  504 (20.7%)  419 (17.2%)  383 (15.7%)  443 (18.2%)  435 (17.9%) | 116 (0.4%)  3,011 (10.7%)  4,154 (14.7%)  3,330 (11.8%)  1,552 (5.5%)  1,208 (4.3%)  14,859 (52.6%) |
| Comorbidity  Congestive Heart Failure  Cancer (Local)  Diabetes Mellitus w/o Complications  Diabetes Mellitus w/ Complications  Dementia  HIV/AIDS  Hypertension  Liver Disease - Mild  Liver Disease - Moderate/Severe  Myocardial Infarction  Cancer (Metastatic)  Peptic Ulcer Disease  Peripheral Vascular Disease  Paraplegia/Hemiplegia  Chronic Pulmonary disease  Renal Disease  Connective Tissue Disease  Cerebrovascular Disease | 248 (10.2%)  372 (15.3%)  381 (15.6%)  295 (12.1%)  20 (0.8%)  6 (0.2%)  1,353 (55.5%)  255 (10.5%)  19 (0.8%)  144 (5.9%)  73 (3.0%)  48 (2.0%)  375 (15.4%)  23 (0.9%)  635 (26.1%)  369 (15.1%)  149 (6.1%)  223 (9.2%) | 707 (2.5%)  1,138 (4.0%)  1,034 (3.7%)  694 (2.5%)  155 (0.5%)  17 (0.1%)  4,878 (17.3%)  911 (3.2%)  91 (0.3%)  406 (1.4%)  263 (0.9%)  261 (0.9%)  1,062 (3.8%)  107 (0.4%)  3,085 (10.9%)  1,027 (3.6%)  434 (1.5%)  759 (2.7%) |
| Immunosuppressant use | 165 (6.8%) | 354 (1.3%) |

**eTable 3: Kaplan-Meier statistical significance tests for hospitalization, ICU admission, and mortality status.**  For each clinical endpoint, we compare the rates in the Bamlanivimab and propensity-matched control cohorts over the course of the study, and we report the Kaplan-Meier log-rank test p-values. This statistical significance test makes the proportional hazards assumption and takes into account the fact that the clinical endpoint is recorded at multiple points in time.  Here, we consider the following clinical endpoints: **(1) Hospitalization:** Whether or not the patient was admitted to the hospital, **(2) ICU admission:** Whether or not the patient was admitted to the ICU, and **(3) Mortality:** Mortality status of the patient.

| **Clinical Endpoint** | **Log-rank test p-value** |
| --- | --- |
| Hospitalization | 0.02 |
| ICU admission | 0.11 |
| Mortality | 0.09 |

**eTable 4: Patient counts for top-20 immunosuppressive medications.**  For each medication, we provide the number of patients in the overall study population (pre-matching) that has received the specified immunosuppressant medication in the past 1 year.  The full list of immunosuppressive medications used to query the EHR database included 819 medications under the WHO ATC L04A.^1^

| **Medication Name Description** | **Patient Count** |
| --- | --- |
| TACROLIMUS 1 MG CAPSULE  TACROLIMUS 0.5 MG CAPSULE  MYCOPHENOLATE MOFETIL 250 MG CAPSULE  METHOTREXATE SODIUM 2.5 MG TABLET  MYCOPHENOLATE MOFETIL 500 MG TABLET  AZATHIOPRINE 50 MG TABLET  MYCOPHENOLATE MOFETIL 200 MG/ML ORAL SUSPENSION  SIROLIMUS 1 MG TABLET  MYCOPHENOLATE SODIUM 180 MG TABLET,DELAYED RELEASE  ADALIMUMAB (CITRATE FREE) 40 MG/0.4 ML SUBCUTANEOUS PEN KIT  MYCOPHENOLATE SODIUM 360 MG TABLET,DELAYED RELEASE  TACROLIMUS 5 MG CAPSULE  ALEMTUZUMAB 30 MG/ML INTRAVENOUS SOLUTION  SIROLIMUS 0.5 MG TABLET  TACROLIMUS 0.2 MG ORAL GRANULES IN PACKET  TOCILIZUMAB IVPB 100 ML (RESTRICTED)  LEFLUNOMIDE 20 MG TABLET  TACROLIMUS 1 MG ORAL GRANULES IN PACKET  HUMIRA PEN (CITRATE FREE) 40 MG/0.4 ML SUBCUTANEOUS KIT  HUMIRA PEN 40 MG/0.8 ML SUBCUTANEOUS KIT | 223  180  157  107  96  52  37  32  28  26  26  26  23  23  22  22  21  20  17  16 |

**eTable 5: Comparison of negative control outcome (cancer screens) for unmatched and matched cohorts.**  Patients with cancer screens were identified by ICD-10 codes (Z12.*) recorded during 2-6 weeks following PCR diagnosis date.  Patients with PCR diagnosis dates after February 8, 2021 were excluded from the analysis due to lack of 42-day follow-up data. The first row shows the comparison of the cancer screening rates between the unmatched cohorts, and the second row shows the comparison between the propensity-matched cohorts. The logistic odds ratio is the coefficient of the treatment variable for a logistic regression model predicting cancer screening rate, controlling for residual confounding factors such as demographics, BMI, comorbidities, and immunosuppressant use. The last column shows the p-value from the Fisher exact test comparing the cancer screening rates between the treated and untreated groups, without controlling for any residual confounding factors.

|  | **Cancer screening rate among treated individuals with follow-up data** | **Cancer screening rate among untreated individuals with follow-up data** | **Logistic odds ratio**  (95% CI) | **Fisher exact test p-value** |
| --- | --- | --- | --- | --- |
| **Unmatched** | 23/2,341 (0.98%) | 129/27,575 (0.47%) | 1.02 (0.64, 1.64) | 0.002 |
| **Matched** | 23/2,251 (1.0%) | 27/2,256 (1.2%) | 1.25 (0.67, 2.31) | 0.48 |

**eTable 6: Intent to treat analysis. Sensitivity analysis including patients who refused bamlanivimab treatment into the bamlanivimab cohort.**

| **Outcome** | **Bamlanivimab cohort**  **(intended)**  (2509 patients) | **Control cohort**  (2509 patients) | **Risk difference**  (95% CI) | **Odds ratio**  (95% CI) | **BH-adjusted p-value** |
| --- | --- | --- | --- | --- | --- |
| Number of patients with follow-up data | | | | | |
| 14 day | 2279 | 2295 |  |  |  |
| 21 day | 2130 | 2139 |  |  |  |
| 28 day | 1927 | 1962 |  |  |  |
| Hospital admission rate | | | | | |
| 14 day | 49/2279 (2.2%) | 84/2295 (3.9%) | 1.5% (0.54%, 2.5%) | 0.57 (0.40, 0.83) | 0.01* |
| 21 day | 57/2130 (2.7%) | 87/2134 (4.3%) | 1.4% (0.31%, 2.5%) | 0.65 (0.45, 0.92) | 0.02* |
| 28 day | 63/1927 (3.3%) | 86/1967 (4.4%) | 1.1% (-0.091%, 2.3%) | 0.77 (0.55, 1.09) | 0.14 |
| ICU admission rate | | | | | |
| 14 day | 8 (0.35%) | 24 (1%) | 0.69% (0.21%, 1.2%) | 0.3 (0.13, 0.70) | 0.02* |
| 21 day | 11 (0.52%) | 24 (1.1%) | 0.61% (0.065%, 1.1%) | 0.43 (0.20, 0.90) | 0.04* |
| 28 day | 17 (0.88%) | 26 (1.3%) | 0.44% (-0.21%, 1.1%) | 0.68 (0.36, 1.29) | 0.23 |
| Mortality rate | | | | | |
| 14 day | 2 (0.088%) | 3 (0.13%) | 0.043% (-0.15%, 0.23%) | 0.79 (0.06, 9.71) | 0.86 |
| 21 day | 5 (0.23%) | 4 (0.19%) | -0.048% (-0.32%, 0.23%) | 1.44 (0.33, 6.21) | 0.86 |
| 28 day | 7 (0.36%) | 6 (0.31%) | -0.057% (-0.42%, 0.31%) | 1.14 (0.31, 4.18) | 0.86 |

**eFigure 6: Rejection probability of relative risk confidence interval-based test and Fisher exact test at 5% significance**. We do 10,000 simulated n=2,000 samples for a proportion *p_2_* (the “control proportion”) ranging from 0.01% to 2%, run the two significance tests on each simulated sample, and compute the proportion of the simulations which reject the test at a 5% significance level. **(A)** shows the rejection probability under the null hypothesis that the relative risk is 1, i.e. the “treatment proportion” *p_1_* = *p_2_*; a dotted line is shown at the nominal significance level of 5%. **(B)** shows the rejection probability under the particular alternate *p_1_ = 0.5p_2_,* and **(C)** shows the rejection probability for *p_1_* = *2p_2_*. Indeed for small *p_2_* there is some range over which the relative risk confidence interval-based test is less powerful than the Fisher exact test; we may be observing this in our study.

**(A)**

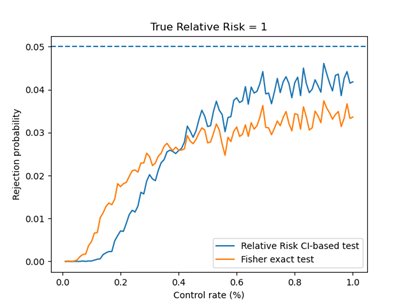

**(B)** **(C)**

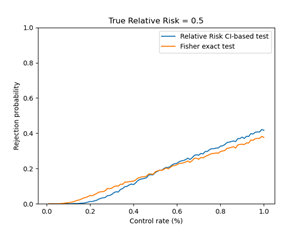

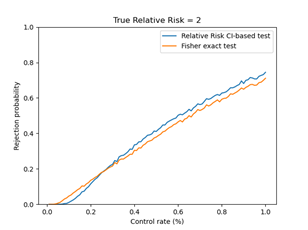

**eSupplement: MATRx team members**

Nicole C. E. Aloia, M.A., M.H.A.; Ryan J. Anderson, Pharm.D.; Gokhan Anil, M.D.; Sara E. Ausman, Pharm.D.; Marcie L. Billings, M.D.; Rachel K. Bishop, B.S.N.; Carl H. Cramer, M.D.; Tracy L. Culbertson, M.S.N.; Ala S. Dababneh, M.D.; Amber N. Derr, M.B.A; Kevin Epps, Pharm.D.; Susan M. Flaker, Pharm.D.; Mary A. Gilmer, Pharm.D.; Eric Gomez Urena, M.D.; Christopher R. Gulden, M.A.; Tamara L. Haack; Jenna R. Herzog; Lex D. Hokanson, D.N.P.; Laura H. Hopkins, M.S.N.; Richard J. Horecki, M.D.; Bipinchandra Hirisave Krishna, M.D.; W. Charles Huskins, M.D., M.Sc.; Tammy A. Jackson, B.A.; Ryan R. Johnson; Betty Jorgenson, M.S.N.; Cory Kudrna; Brian D. Kennedy, Pharm.D.; Mary K. Klingsporn, M.S.N.; Brian Kottke, M.B.A; Sarah R. Lessard, Pharm.D.; Larry I. Lutwick, M.D.; Edward J. Malone III, M.D.; Jennifer A. Matoush, APRN, CNS, M.S.; Ivana N. Micallef, M.D.; Darcie E. Moehnke, M.A.N.; Muhanad Mohamed M.B.B.S.; Colleena N. Ness; Shelly M. Olson, M.S.N.; Raj Palraj, M.B.B.S.; Janki Patel, D.O.; Damian J. Paulson; David Phelan, M.D.; Margaret T. Peinovich, Pharm.D.; Wilford L. Ramsey, M.H.A.; Taunya J. Rau-Kane; Kevin I. Reid, D.M.D.; Karen J. Reinschmidt, M.S.; Maria Teresa Seville, M.D.; Erin C. Skold, J.D.; Jill M. Smith, APRN; Laurie A. Spielman, M.S.N.; Donna J. Springer, APRN, CNS, M.S.; Perry W. Sweeten, Pharm.D.; Jennifer M. Tempelis, Pharm.D.; Paschalis Vergidis, M.D.; Daniel C Whipple, M.S.
